# The accuracy of machine learning models using ultrasound images in prostate cancer diagnosis: A systematic review

**DOI:** 10.1101/2022.02.03.22270377

**Authors:** Retta C Sihotang, Claudio Agustino, Ficky Huang, Dyandra Parikesit, Fakhri Rahman, Agus Rizal AH Hamid

## Abstract

Prostate Cancer (PCa) is the third most commonly diagnosed cancer worldwide, and its diagnosis requires many medical examinations, including imaging. Ultrasound offers a practical and cost-effective method for prostate imaging due to its real-time availability at the bedside. Nowadays, various Artificial Intelligence (AI) models, including Machine learning (ML) with neural networks, have been developed to make an accurate diagnosis. In PCa diagnosis, there have been many developed models of ML and the model algorithm using ultrasound images shows good accuracy. This study aims to analyse the accuracy of neural network machine learning models in prostate cancer diagnosis using ultrasound images. The protocol was registered with PROSPERO registration number CRD42021277309. Three reviewers independently conduct a literature search in five online databases (MEDLINE, EBSCO, Proquest, Sciencedirect, and Scopus). We screened a total of 132 titles and abstracts that meet our inclusion and exclusion criteria. We included articles published in English, using human subjects, using neural networks machine learning models, and using prostate biopsy as a standard diagnosis. Non relevant studies and review articles were excluded. After screening, we found six articles relevant to our study. Risk of bias analysis was conducted using QUADAS-2 tool. Of the six articles, four articles used Artificial Neural Network (ANN), one article used Recurrent Neural Network (RNN), and one article used Deep Learning (DL). All articles suggest a positive result of ultrasound in the diagnosis of prostate cancer with a varied ROC curve of 0.76-0.98. Several factors affect AI accuracy, including the model of AI, mode and type of transrectal sonography, Gleason grading, and PSA level. Although there was only limited and low-moderate quality evidence, we managed to analyse the predominant findings comprehensively. In conclusion, machine learning with neural network models is a potential technology in prostate cancer diagnosis that could provide instant information for further workup with relatively high accuracy above 70% of sensitivity/specificity and above 0.5 of ROC-AUC value. Image-based machine learning models would be helpful for doctors to decide whether or not to perform a prostate biopsy.

## Introduction

Prostate cancer (PCa) is the third most common cancer globally and the second most common cancer in men. In 2020, there were an estimated 1,4 million new cases and 375,000 new prostate cancer deaths worldwide.[1] There are several modalities to diagnose PCa, including digital rectal examination (DRE), prostate-specific antigen (PSA) levels, biomarkers, imaging, and histopathology. The current gold standard for PCa detection in core needle biopsy, performed under transrectal ultrasound (TRUS) guidance.[2-5] The role of ultrasound (US) in this procedure is not for targeting PCa but for anatomical navigation. Aside from the complications associated with the biopsy, high levels of underdiagnosed and overtreatment have been reported.[6-7]

Ultrasound is a potential candidate for PCa imaging because it is cost-effective, practical, and widely available. The problem of ultrasound images interpretation is that hypoechoic areas suspected of cancer can be normal or cancerous histologically. Most prostate cancers are hypoechoic on TRUS, whereas 30%-40% of prostate cancer are isoechoic, and 1.5% are hyperechoic.[8] The sensitivity and specificity of TRUS are limited, ranging between 40% and 50% for detecting PCa.[8,9] Several studies depict that using grey mode US alone is inadequate for PCa screening.[10] However, there are several new modes of US in prostate gland imaging, such as contrast-enhanced ultrasound (CEUS), color Doppler and ultrasound elastography. CEUS is an ultrasound technique that uses intravenous injected gas-filled microbubbles as a contrast agent to provide microvascular and tissue perfusion information.[11] Color Doppler mode has capability to detect motion or blood flow using a color map to show the speed and direction of blood flow through vessel.[12] Ultrasound elastography is a non-invasive imaging technique to measure changes in soft tissue elasticity.[13] Those new modes increase overall accuracy in detecting PCa in comparison to grey mode.[10-17] However, the diagnostic performance of those modes is still not quite satisfactory with a wide range of sensitivity (67% to 93%) and specificity (59% to 93%).[12-13,16-17]

Artificial Intelligence (AI) is defined as an ability of a computer to perceive the surrounding environment and make the same decisions as a human intellect on an action to reach a particular goal.[18] AI is now a revolutionising technology in the healthcare field and is gaining interest. Machine learning (ML) is a branch of AI that focuses on using data and algorithms to improve accuracy. To train machine learning, it is enough to acquire structured datasets consisting of input variables and outcomes. AI has a vital role in interpreting large amounts of data. Neural networks (such as artificial neural networks – ANN, convolutional neural networks – CNN, recurrent neural networks – RNN) are machine learning models which work like human biological neuron. They have ability to learn and model non-linear and complex relationships that enable them to generate relationships between inputs and outputs in a complex pattern. The developed algorithm of machine learning may help urologists to reduce the number of unnecessary prostate biopsies without missing the diagnosis of aggressive PCa.[19]

In PCa, AI has been shown to aid in a standardised pathological grading to assess cancer stratification and treatment. Numerous studies have evaluated the utility of prostate specific antigen (PSA) and/or magnetic resonance imaging (MRI) in the setting of AI in detecting prostate cancer. Nitta et al.[20] and Djavan et al.[21] applied machine learning models to predict prostate cancer based on PSA levels. Machine learning tends to be superior to the conventional methods with ROC-AUC ranging from 0.63-0.91 based on the machine learning models and PSA categories. Aldoj et al.[22] utilized AI using MRI with 3D combinations (apparent diffusing coefficient (ADC), diffusion weighted imaging (DWI), and T2 weighted images) which generated sensitivity at 81.2% and specificity at 90.5%. The high diagnostic performance is also found in a study by Yoo et al.[23] using deep CNN analysis of DWI sequence with ROC-AUC of 0.84-0.87. However, the accuracy of machine learning using ultrasound data as the primary modality remains debatable. This review aims to analyse the accuracy of neural networks ML models using ultrasound images in prostate cancer diagnosis.

## Methods

### Protocol Registration

The protocol for this systematic review was registered with PROSPERO registration number CRD42021277309.

### Search Strategy

Three reviewers independently (RC, CA, FH) conducted a literature search in five online databases (MEDLINE, EBSCO, Proquest, ScienceDirect, and Scopus) on November 19^th^, 2021. The keywords used were “Prostate Cancer” AND “Machine Learning” AND “Diagnosis” AND “Ultrasonography with various combinations as written in **Table 1**. The researcher also reviewed the reference list of chosen articles from the literature search to identify relevant studies.

**Table 1.** Literature search strategy

| Database | Keywords | Result | Date attempt |
| --- | --- | --- | --- |
| PubMed | ((((prostate cancer or prostate carcinoma) AND (((imaging) OR (Ultrasonography)) OR (transrectal ultrasonography)) OR (TRUS))) AND ((diagnostic outcome) AND (diagnosis))) AND (((machine learning) OR (deep learning)) OR (artificial neural network)) OR (convolutional neural network)) | 78 | November 19<sup>th</sup>, 2021 |
| EBSCO | machine learning or artificial intelligence or deep learning or neural network AND prostate cancer AND ultrasound or sonography or ultrasonography<br>[Title] | 7 | November 19<sup>th</sup>, 2021 |
| Proquest | ab(prostate cancer) AND ab(ultrasound) AND ab(artificial neural network OR machine learning) AND ab(diagnosis) | 4 | November 19<sup>th</sup>, 2021 |
| Sciencedirect | Title, abstract, keywords: (prostate cancer) AND (ultrasound) AND (machine learning OR neural network) AND (diagnosis)<br>[research articles] | 13 | November 19<sup>th</sup>, 2021 |
| Scopus | TITLE-ABS-KEY ( machine AND learning AND ultrasound AND prostate AND cancer AND diagnosis ) | 43 | November 19<sup>th</sup>, 2021 |

### Study Selection and Data Extraction

We included all available articles about machine learning use in prostate cancer diagnosis that used ultrasound images. We restricted our search to articles published in English without a publication date limit. A study was considered relevant if it fulfilled our inclusion criteria: using human subjects, using neural networks machine learning models, and using prostate biopsy as a standard diagnosis. We included cohort, case-control, and cross-sectional studies. An article was excluded from the selection if it was a conference/review article, combination examination with MRI, or no diagnostic parameter in the article. All reviewers screened the title and abstract of the selected papers independently. Discrepancies between the reviewers were solved through discussion with the senior reviewers (DP, ARAH, and FR) until the consensus was reached. All authors agreed on the final list of selected papers for extraction. The PRISMA flow diagram is used to guide the article selection process.

### Risk of Bias Assessment

Three reviewers independently evaluated the methodological quality of the studies using the QUADAS-2 Tool in Review Manager software 5.4 version. The reviewers were not blinded for the author, journal, or publication identities of each article. The risk of bias assessment consists of three categories: high, unclear, or low risk of bias based on the pre-listed questions in the QUADAS-2 Tools.

## Results

Our electronic search identified 145 articles and only 6 that met our inclusion and exclusion criteria **(Fig 1)**. Four articles use the artificial neural networks (ANN) method, one article uses the recurrent neural network (RNN) method, and one article uses the deep learning (DL) method. The characteristics of each study are described in Table 2. The six included studies used a cross-sectional study design. All articles studied adult male human subjects with an unknown age range due to the unclear data. The sample size ranges from 61 to 1077 patients; however, a study from Ronco et al.[24] only provided the number of cases.

**Table 2.**
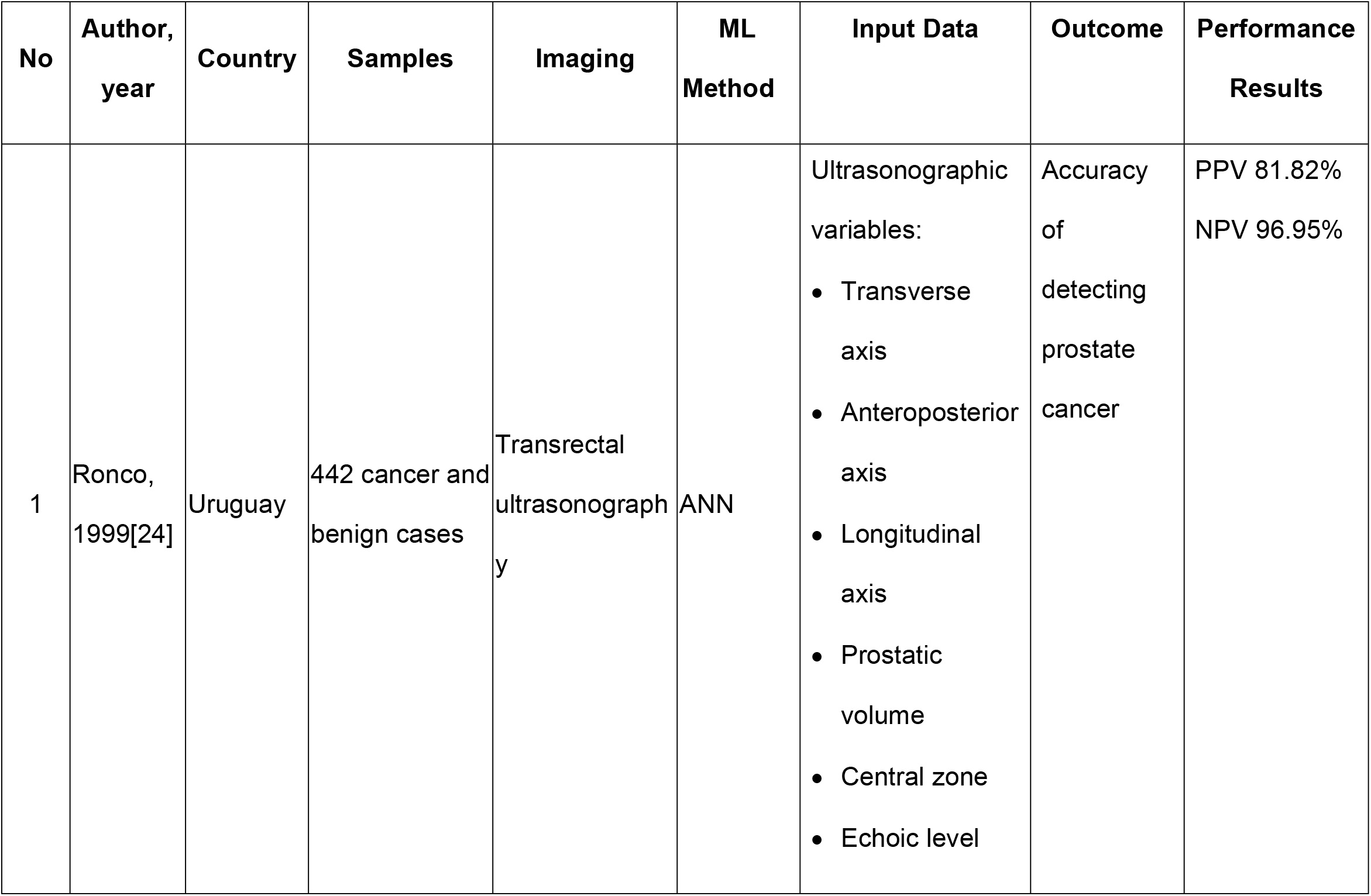
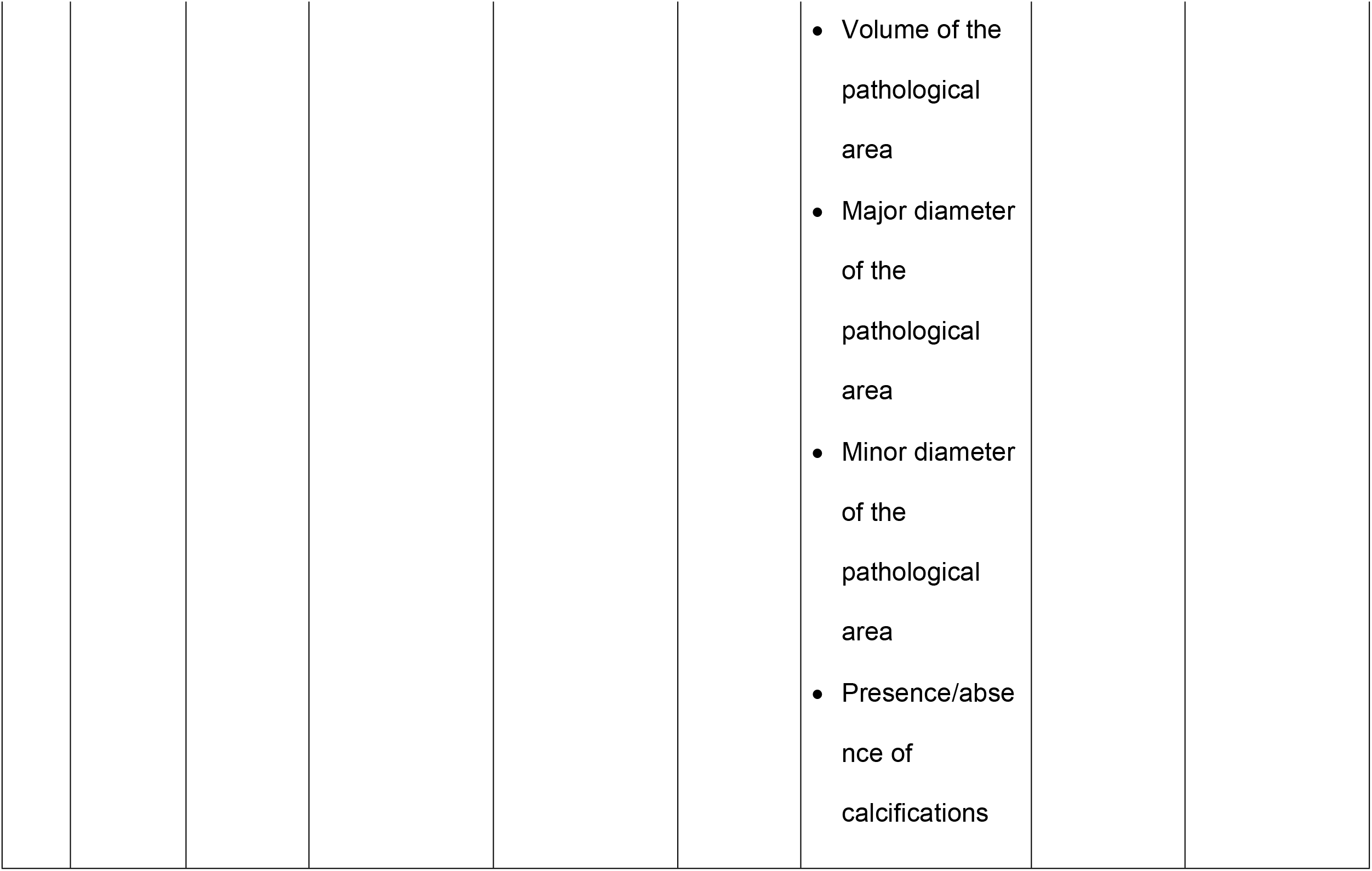
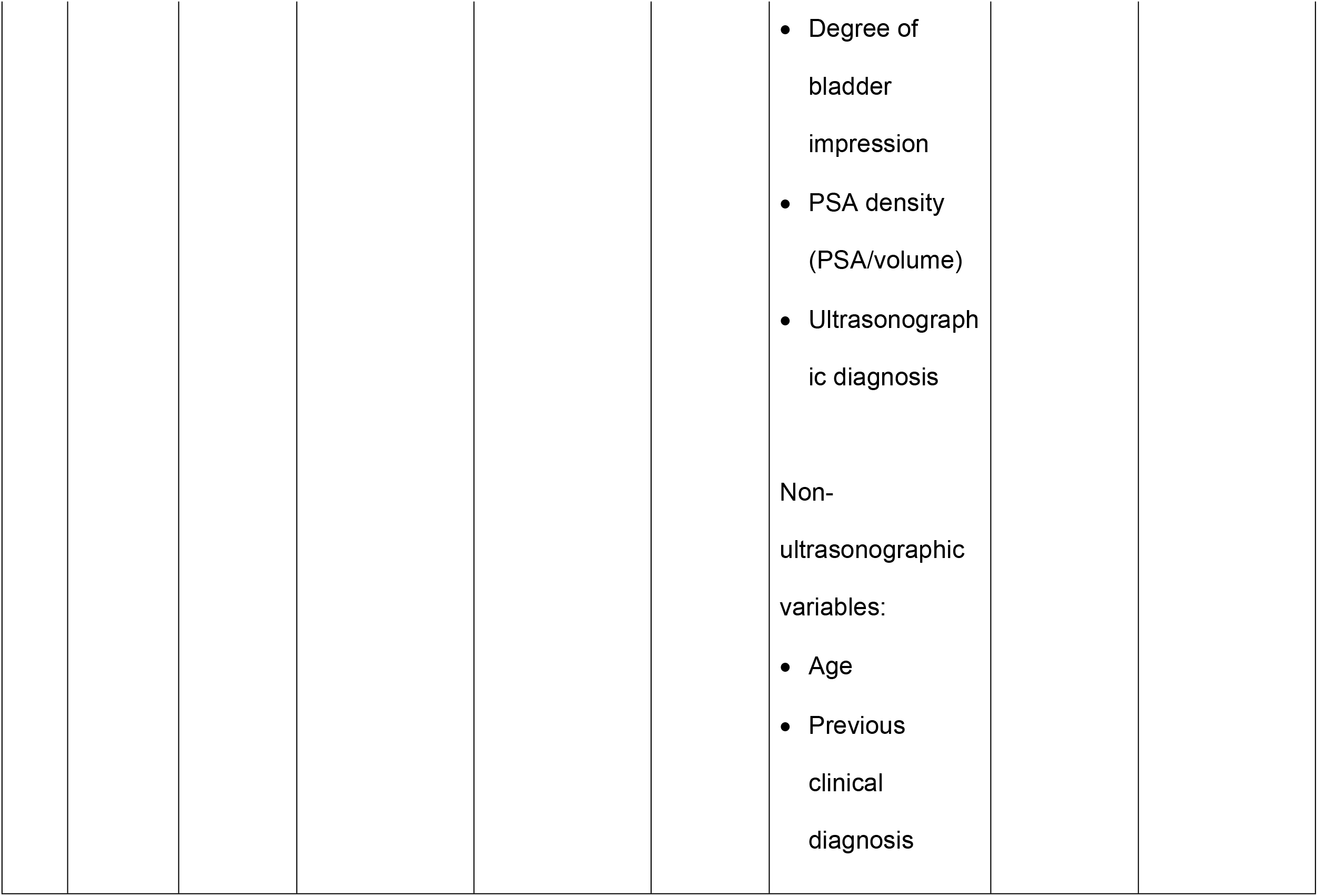
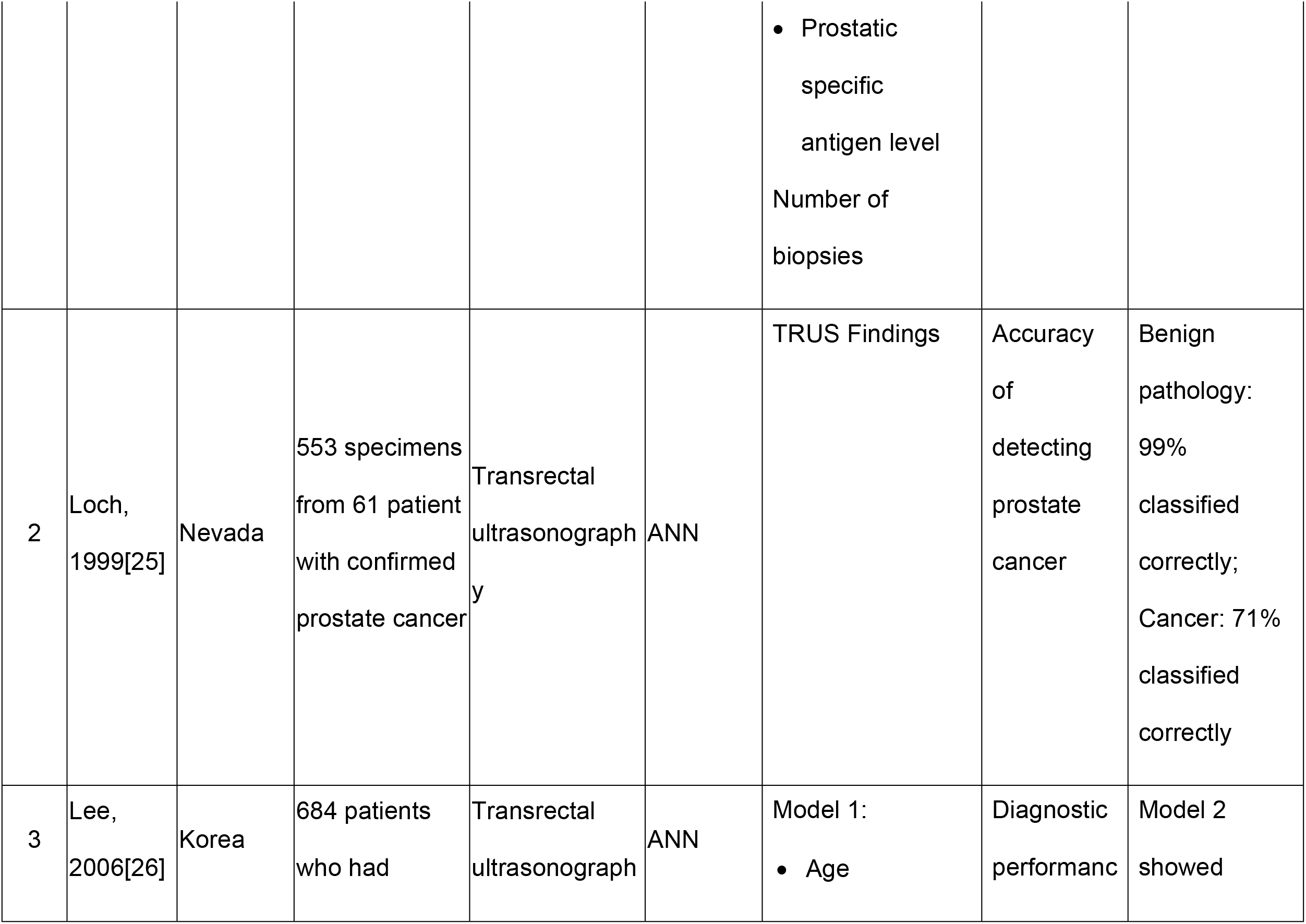
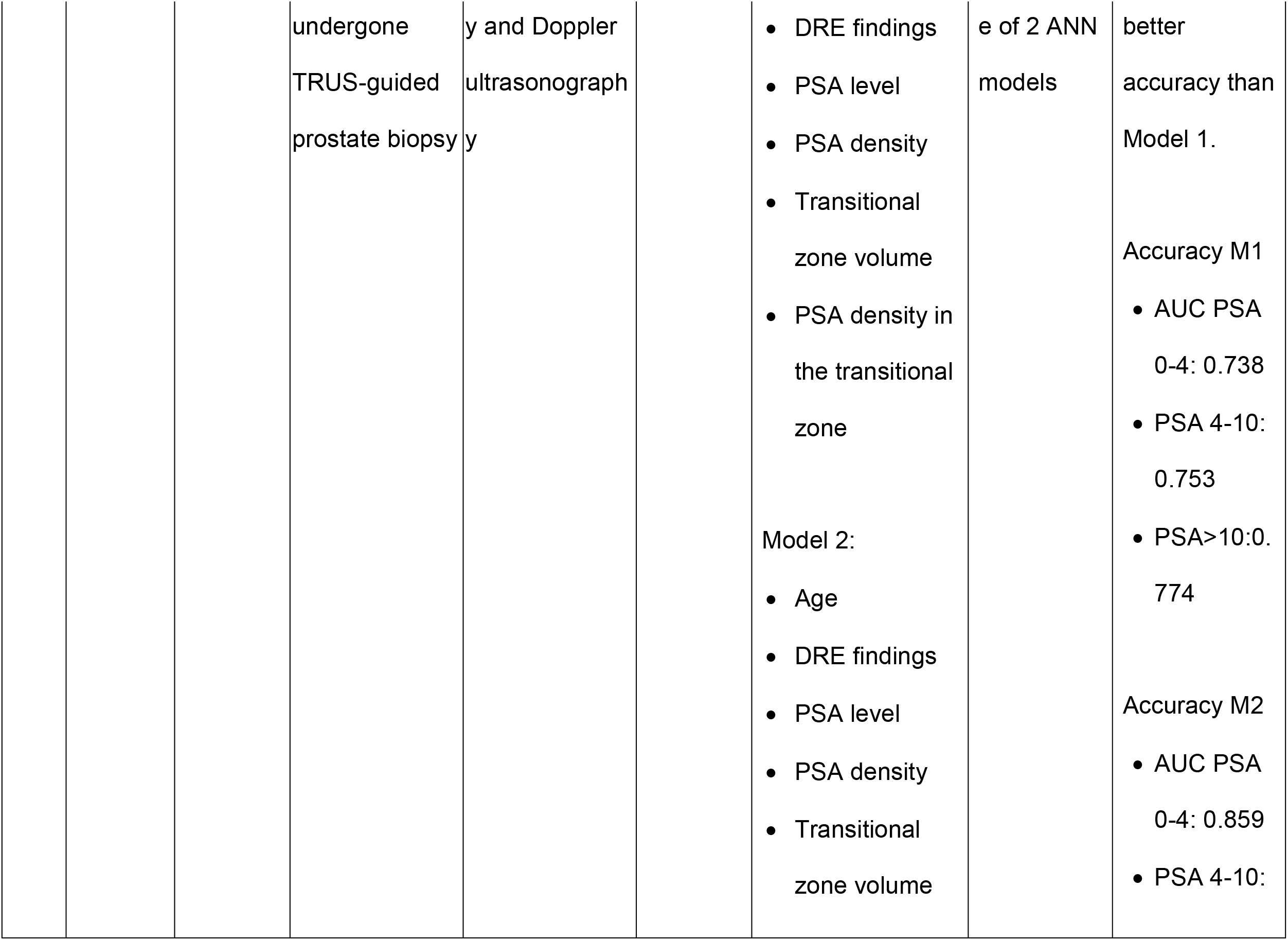
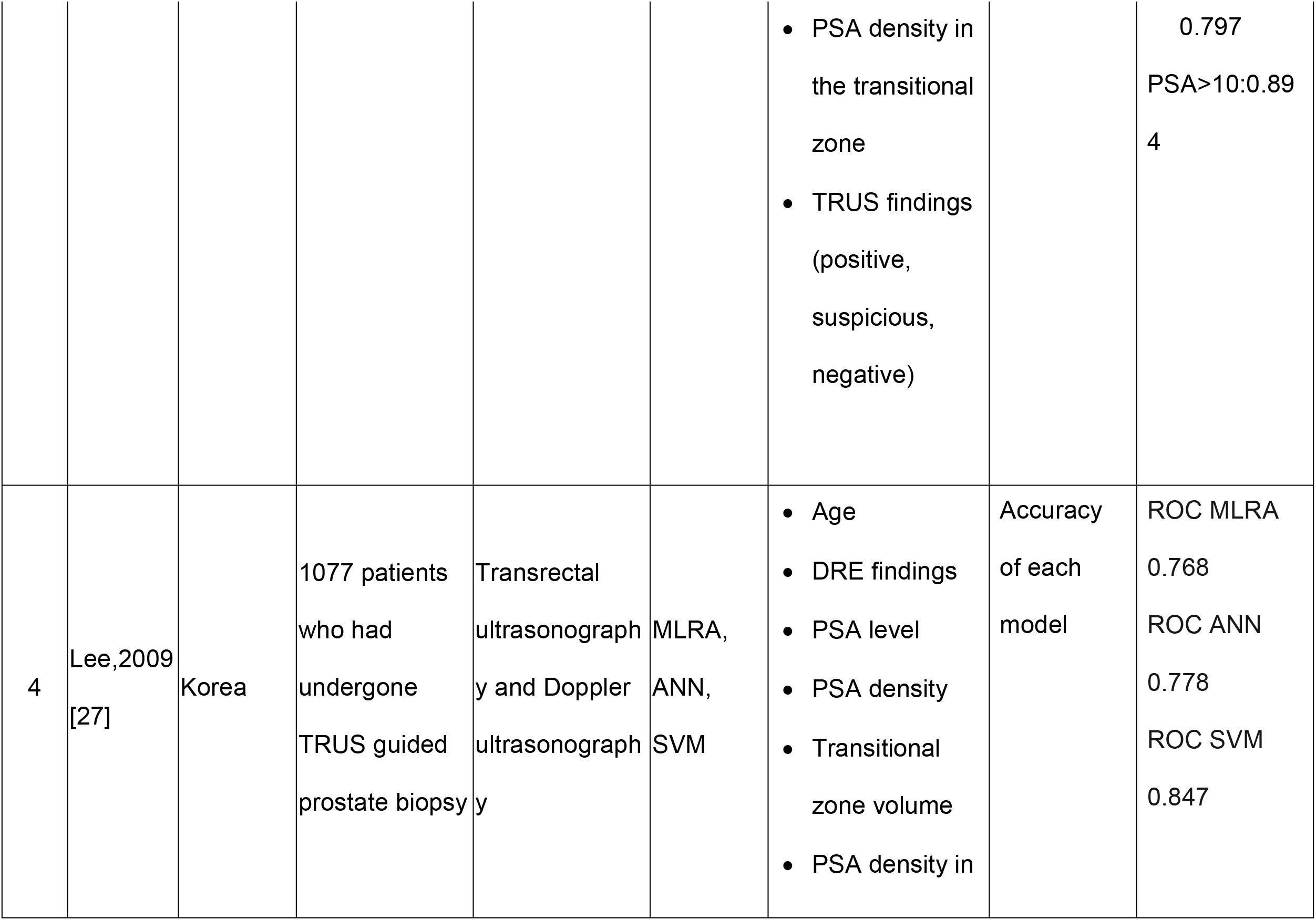
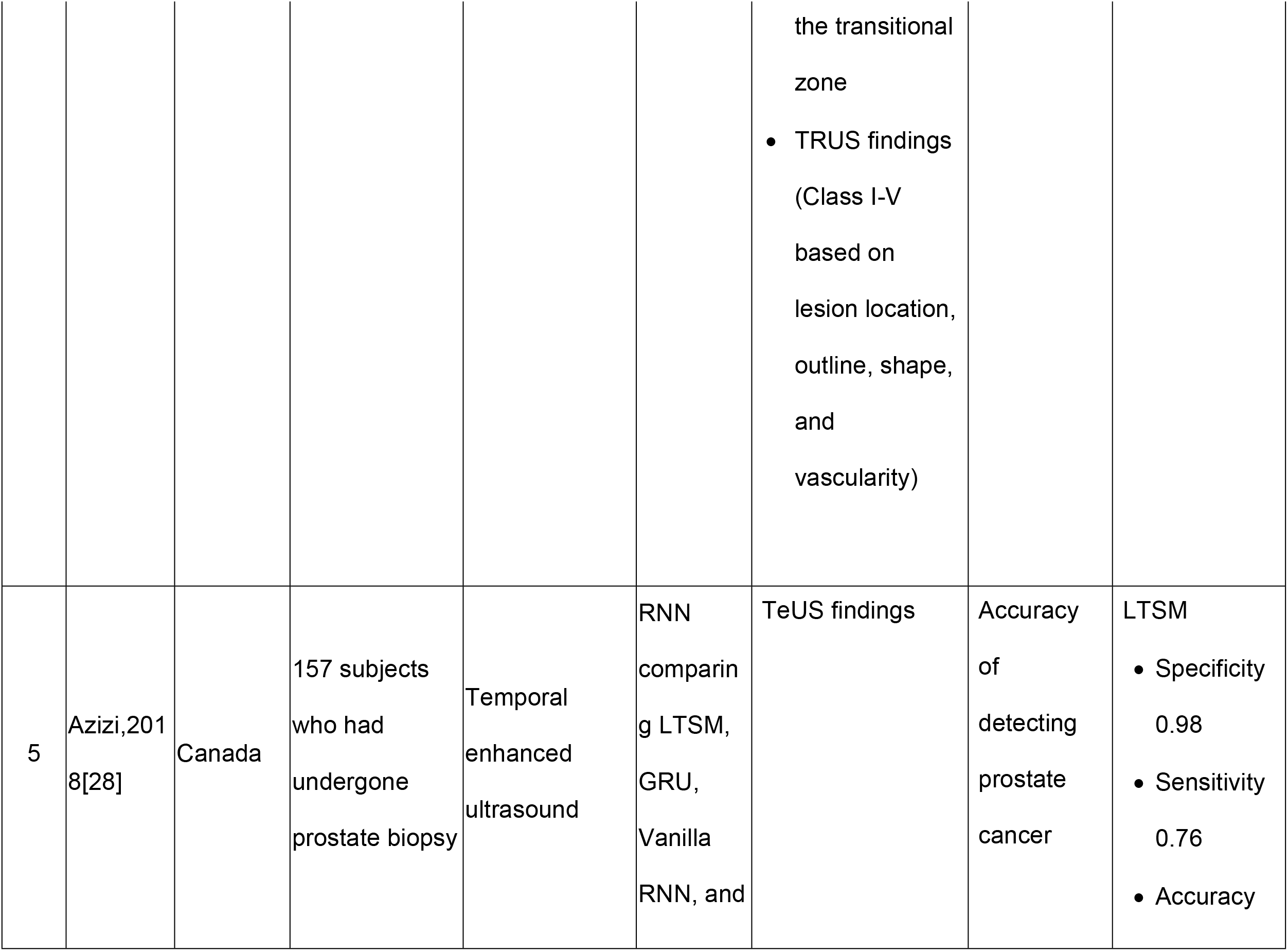
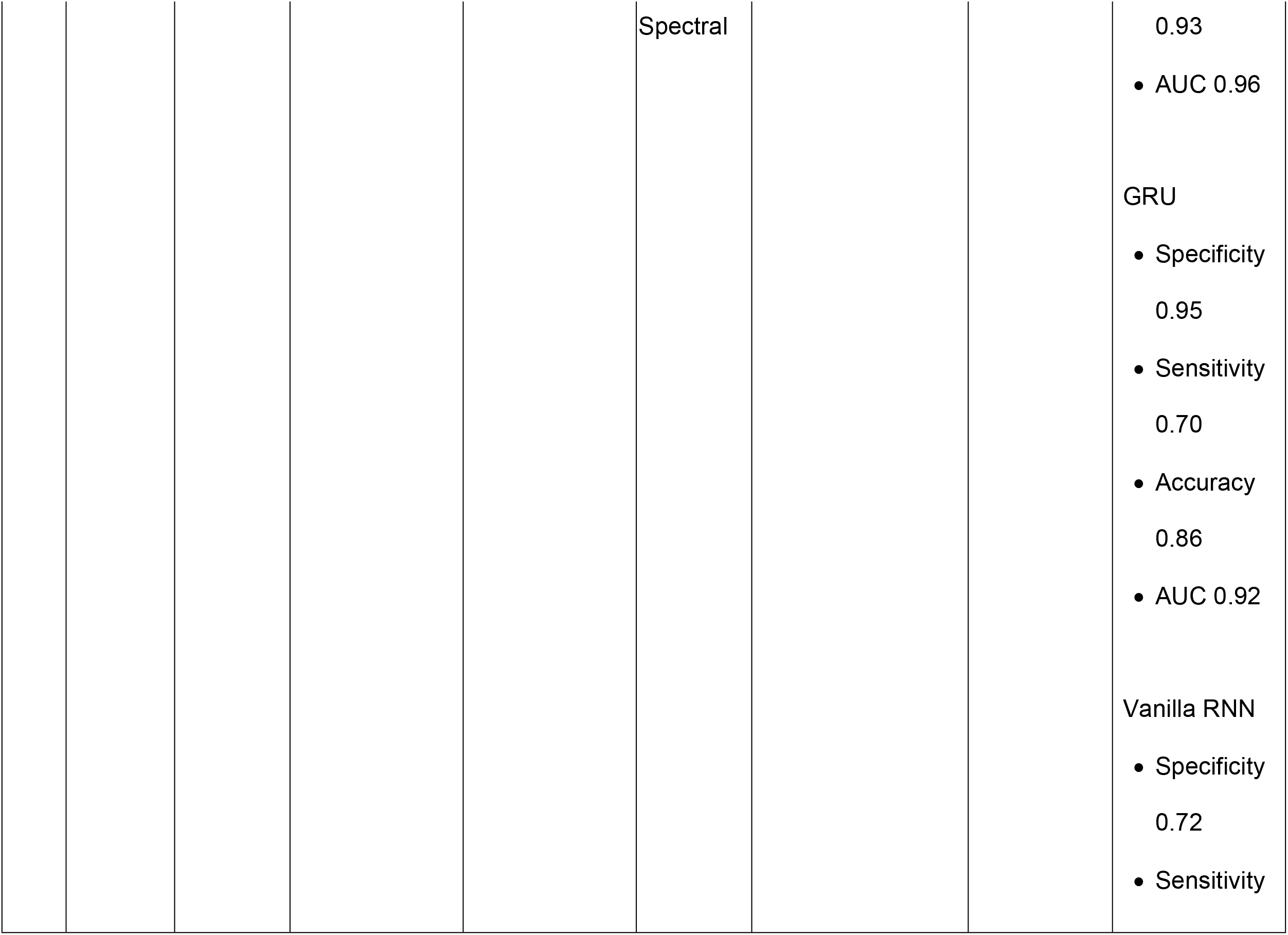
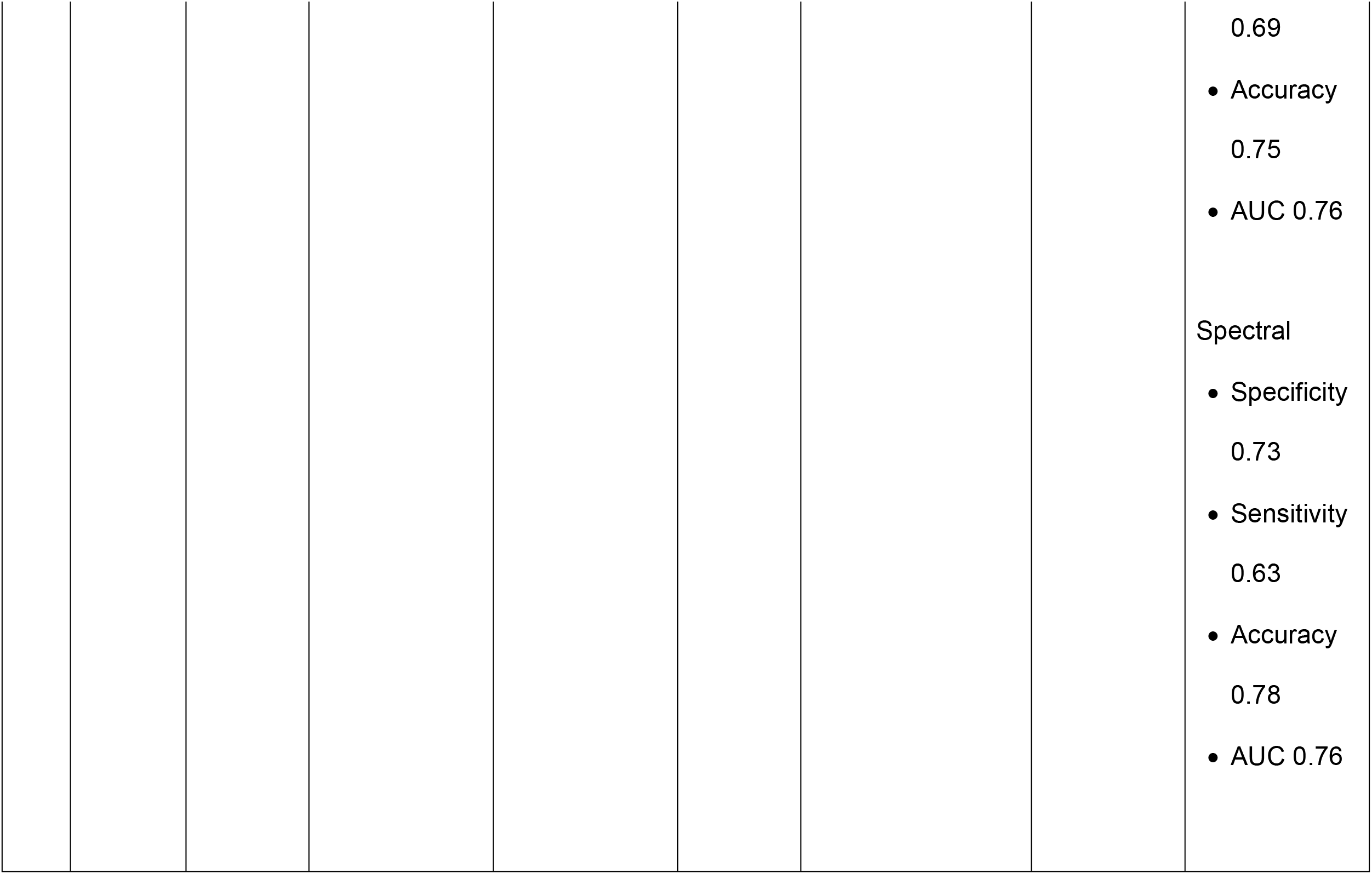
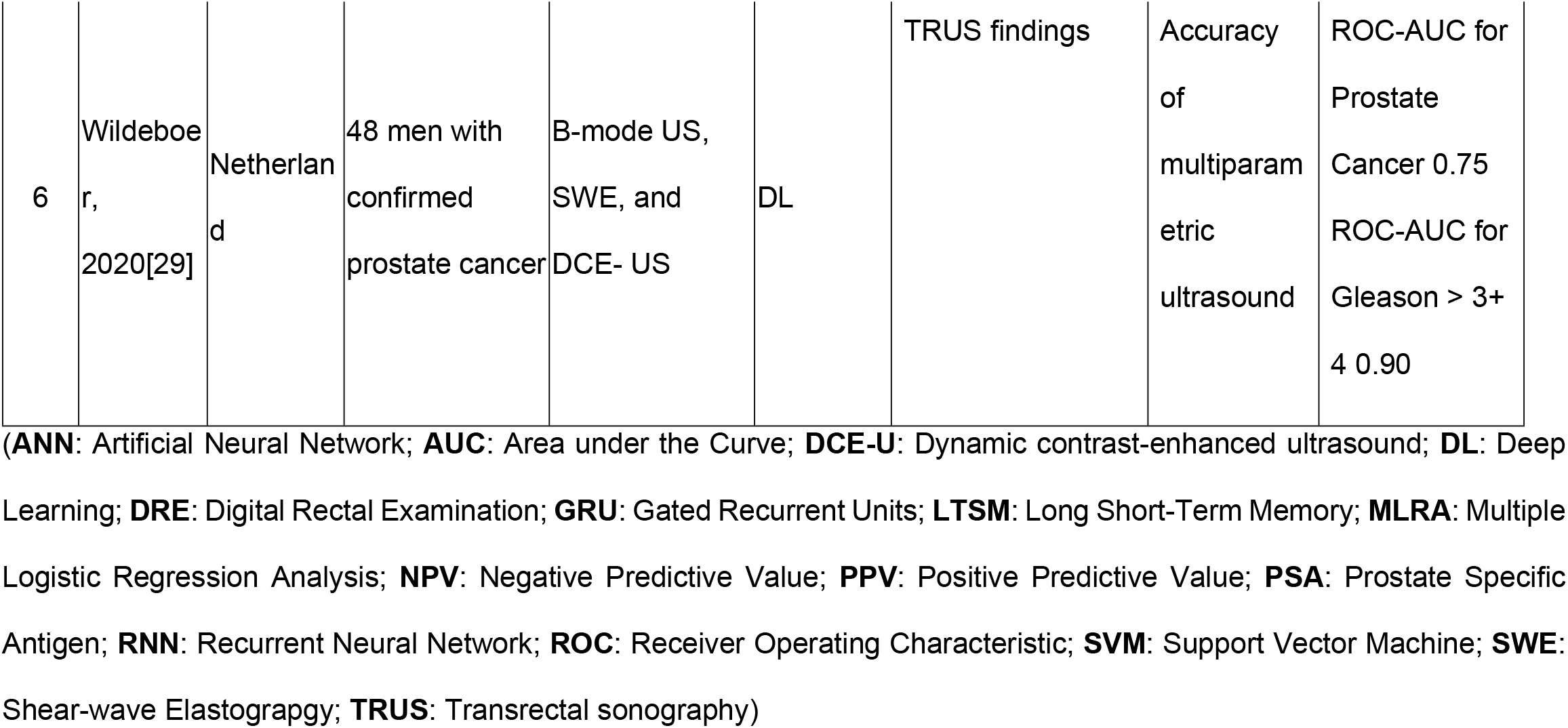
Characteristics and Performance Result of Included Studies

**Figure 1.**
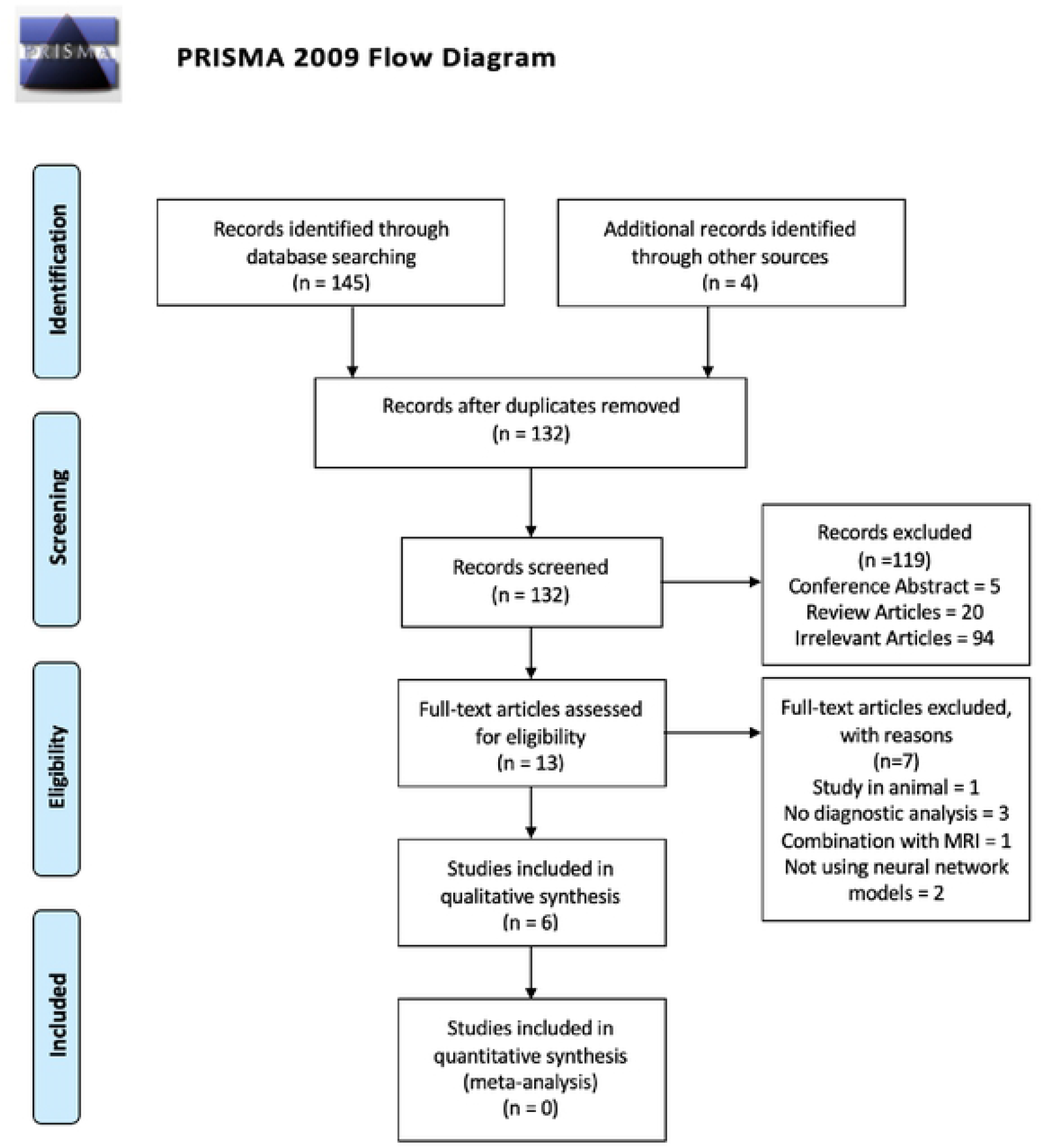
Prisma Flow Diagram

The quality assessment of the six included articles is shown in **Fig 2** using the QUADAS-2 Tool. Several articles have an unclear and high risk of bias. However, we still included the articles in our analysis. The unclear risk of bias is most commonly found in Index Test parameters because of the unclear threshold of the Index test. The high risk of bias is also most commonly found in Index Test parameters due to the results were interpreted with the knowledge of reference standard results in several articles.[24-26]

**Figure 2.**
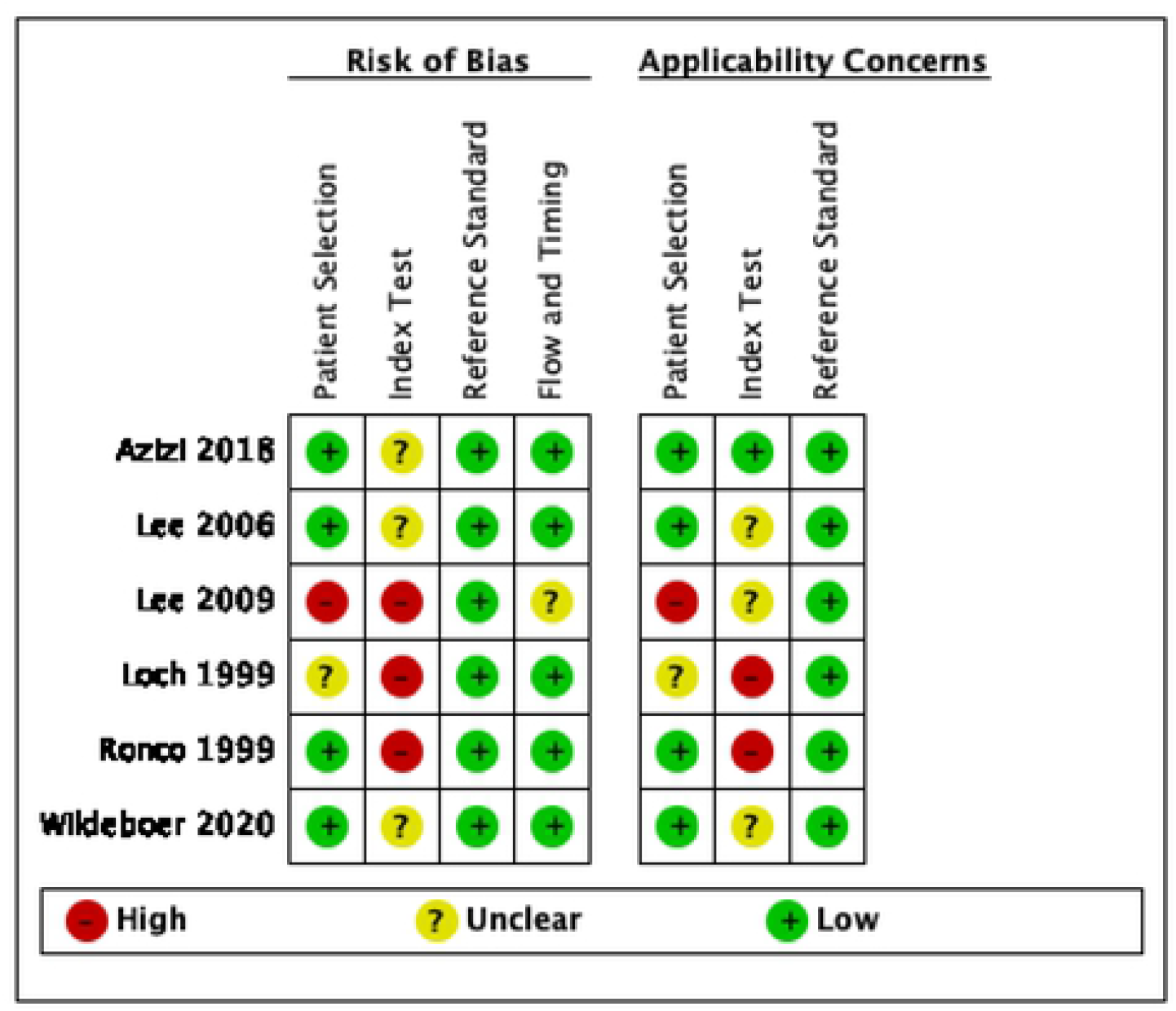
Risk of Bias Assessment using QUADAS-2 Tool

Three articles used TRUS data only for the input parameter and the rest three articles used combination input data from clinical findings. The included studies showed various parameters of accuracy analysis, including ROC-area the curve (AUC), positive predictive value (PPV), negative predictive value (NPV), sensitivity, and specificity **(Table 2)**. However, a study from Loch et al.^25^ used percentage only. The performance results can be seen in **Table 2**. Due to the varied parameters, a quantitative analysis could not be performed. Most of the articles used ROC-AUC as the accuracy parameters.

## Discussion

### Ultrasound in Prostate Cancer

PCa may be suspected when the PSA level increases above normal or if the digital rectal examination (DRE) is abnormal.[2,3] After that, further examinations are carried out to make the diagnosis, including imaging and biopsy as a standard diagnosis. Ultrasound and MRI have been assessed for their ability to reliably detect PCa in men suspected with PCa. MRI allows better visualization of prostate anatomical zones and location of tumor with the extension within the gland. It is useful to allow lesion detection and enable functional imaging of the prostate.[30] The use of MRI as a single modality to detect PCa has been evaluated. Several studies revealed MRI carries a relatively high sensitivity, but poor specificity. Multiparametric MRI has been used widely and shows good sensitivity in larger tumors. However, it is less sensitive to detect lower grade PCa (ISUP Grade 1) with pooled sensitivity of 0.70 and pooled specificity of 0.27.[31] From the current recommendation, multiparametric MRI become the clinical routine examination for patients with suspected PCa and PCa staging.[32-34]

Ultrasound is a cost-effective and widely available imaging modality. However, standard TRUS is not a reliable imaging method due to the low sensitivity and specificity in detecting prostate cancer.[5,9,35] TRUS was initially developed with the aim to guide transperineal biopsies. After being evaluated for years, it became evident that cancers of the prostate were most often anechoic or hypoechoic. However, prostatitis and focal infarct also have been reported to have the appearance of hypoechoic lesions on ultrasound, which cause false positive results.[36]

TRUS has several limitations in basic modes, such as similar backscatter signals of cancerous and normal prostate parenchyma and heterogeneity of the transitional zone.[37] Other modes of TRUS such as color Doppler, CEUS, TeUS, and elastography revealed better performance than B-mode.[29,38-39] Recently, innovations have been made to improve accuracy in detecting prostate cancer. Foster et al. developed ultrasound biomicroscopy, performed at frequencies ranging between 14 and 29 MHz, with theoretical spatial resolution at this frequency being between 50 and 70um. This innovation allowed the visualisation of prostate anatomy details that usually could not be seen in conventional US examinations.[40,41] Despite the limitations, TRUS guided biopsy is still the gold standard for the diagnosis pf prostate cancer.

Although MRI has excellent ability to identify clinically significant PCa, MRI is quite expensive, not portable, and not readily available in healthcare centers. In addition, MRI could not provide real time imaging compared to ultrasound. With such disadvantages, TRUS still has a potential role as imaging modality in prostate cancer diagnosis.[30] A comparative study by Zhang et al showed multiparametric TRUS (grayscale, color Doppler, shear wave elastography, and contrast enhanced ultrasound) had higher sensitivity, negative predictive value, and accuracy than multiparametric MRI (T2-weighted, diffusion-weighted, and dynamic contrast-enhanced MRI) in detecting localized PCa.[42] From evidences, TRUS has some advantages in detecting localized PCa from MRI with lower cost, real time, and higher availability.

### Machine Learning increasing the Role of TRUS in Prostate Cancer Diagnosis

Ultrasound imaging is limited by operator dependence and poor reproducibility. To read ultrasound images, it requires years of experience and training. To overcome the limitations, machine learning have been developed in medical imaging to accelerate ultrasound image analysis and generate an objective disease classification.[43] In recent years, applications of ML to US is developing and rapidly progressing. ML can help by decreasing the time of reader that interprets the amount of data to make a conclusion.[44] ML is a subdiscipline of AI where computer programs learn associations of predictive power from examples of data.[45] Several methods such as classification, regression, registration, and segmentation applied to analyse ultrasound images. However, neural networks algorithms have been shown to significantly improve performance when compared to other classifiers.[43] Neural networks, which work like human brain, gives capability to solve problems based on the available data. This model can incorporate many variables and produce results in more complex situations.[45] In PCa diagnosis, ML can generate input data from various variables to classify whether the patient is suspected of having prostate cancer or not **(Fig 3)**.

**Figure 3.**
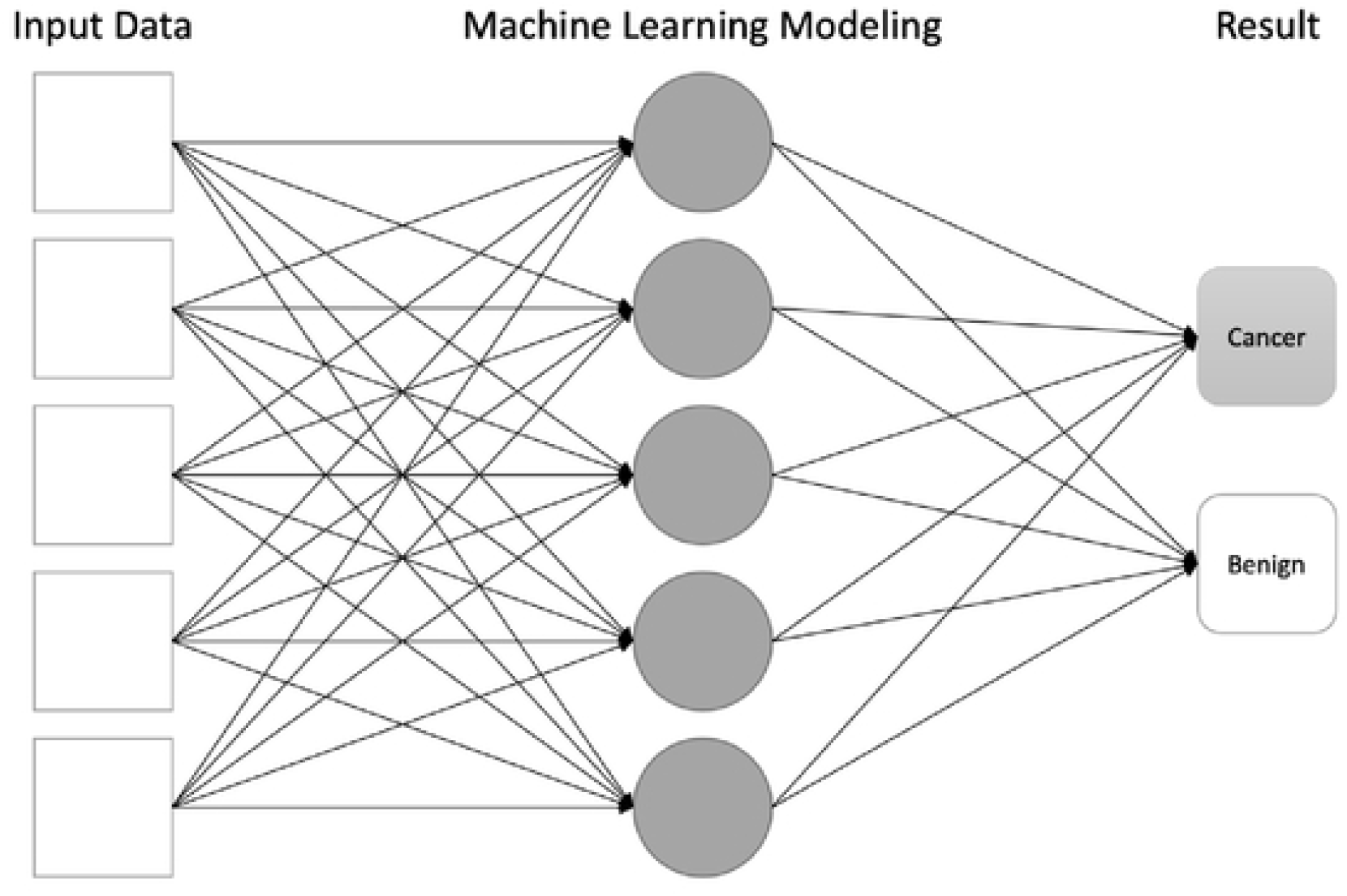
Schematic Machine Learning Model in Prostate Cancer Diagnosis

Input data includes all available variables that could be beneficial to generate a conclusion. Machine learning modeling consists of complex hidden layers which take an essential role in data processing. The result part is a conclusion of the machine learning build-up algorithm based on the input data.

Based on our included studies, the overall accuracy of machine learning shows promising results. ROC-AUC values of five studies showed a number greater than 0.5 with a range of 0.75 to 0.98. Wildeboer et al.[29] assessed the potential DL model based on TRUS’s B-mode, SWE, and DCE-US. The multiparametric classifier reached ROC-AUC of 0.90 compared to 0.75 of the best performing individual parameter for PCa and Gleason >3 + 4 significant PCa. The study revealed that combinations of the available modes were favoured compared to single-mode. Lee et al.[26] evaluated the accuracy of the multiple logistic regression model, ANN model, and support vector machine model to predict prostate biopsy outcomes. The models were constructed from the input data of age, DRE findings, PSA parameters, and TRUS findings. This study showed that image-based clinical decision support systems (ANN and SVM) have better accuracy than the multiple logistic regression model. However, SVM was superior to the performance of both ANN and the multiple logistic regression model. Lee et al.[27] evaluated the diagnostic performance of the ANN model with and without TRUS data. This study included 684 patients who underwent prostate biopsy, with 214 confirmed to have prostate cancer. ANN model was used with primary input data of age, PSA levels, and DRE findings. However, with additional TRUS data, the accuracy of the ANN model was found to be more accurate with a higher value of ROC-AUC. Azizi et al.[28] proposed temporal modeling of TeUS using RNN to improve cancer detection accuracy. TeUS data were acquired from 157 subjects during fusion prostate biopsy. This model achieves a ROC-AUC value of 0.96.

The various levels of accuracy are affected by several factors, including the model of AI, modes of TRUS, amount of input data, Gleason grading, and PSA levels. Modes of TRUS are significantly associated with accuracy, where the DCE-US/SWE/TeUS will improve the visualisation and differentiation of prostate tissues compared to the B-mode. The amount of input data is also an essential factor in making an accurate result in ANN models. More complex data will create a more precise diagnosis.[44] Studies by Lee et al.[27] and Wildeboer et al.[29] revealed that more data combinations would increase the ROC-AUC value, increasing accuracy. Wildeboer et al.[29] showed a significant relationship in Gleason score > 3+4, but no significant result in Gleason score 3+3 or 3+4. This might be due to a bias in patient selection; tumors in 3+3 are considered disproportionately large for clinicians, thus, not included in the study. Based on a study by Lee et al.[27], the ROC-AUC of ANN models is consistently higher in PSA levels above 10. This might be associated with serum PSA levels which correlate with the extent of cancer and histological grade.[46] Thus, TRUS is not reliable to detect PCa as a single tool. However, with utilization of machine learning and combinations of relevant input data, TRUS has a potential role.

### Future Development of Machine Learning-TRUS Model

The machine learning field is advancing rapidly and is supported by new hardware and software technology development. High-resolution and multiparametric imaging can be fused and integrated with other data sets to diagnose prostate cancer better.[47,48] The utilization of machine learning with TRUS data could have a potential role as a diagnostic modality, especially where MRI is not available. Based on the current guidelines, T2-weighted imaging remains the most useful imaging method for local imaging on MRI.[49] However, a meta-analysis by Rooij et al.[50] MRI has high specificity but poor sensitivity for local PCa staging with sensitivity and specificity for extracapsular extension (ECE), seminal vesicle invasion (SVI), and overall stage T3 detection of 0.57 (95% confidence interval [CI] 0.49-0.64) and 0.91 (95% CI 0.88-0.93), 0.58 (95% CI 0.47-0.68) and 0.96 (95% CI 0.95-0.97), and 0.61 (95% CI 0.54-0.67) and 0.88 (95% CI 0.85-0.91), respectively. Our findings showed that machine learning with TRUS other relevant data could increase the diagnostic performance. Thus, it will become more affordable and easier to diagnose PCa by not performing MRI. Furthermore, the use of TRUS with machine learning can be implemented as a fused combination with MRI to do a prostate biopsy and intraoperative mapping to register preoperative MRI during robotic surgery. This feature allows the surgeon to visualize the suspected lesions on the instrument display during the procedure.

## Limitation of the Study

We provide all available evidence about machine learning models of human ultrasound images in prostate cancer diagnosis. However, none of the articles shows the same output parameters to generate a quantitative analysis. Our approach included a comprehensive search of multiple databases as well as other sources for relevant publications. Since we restricted our literature search in English, some articles in other languages may be missed out. The major weakness of this study is low to moderate quality of included studies and the limited number of studies. Although there was only limited evidence, we managed to analyse the predominant findings comprehensively.

## Conclusions

Machine learning with neural network models is a potential technology in prostate cancer that could provide instant information for further workup with relatively high accuracy above 70% of sensitivity/specificity and above 0.5 of ROC-AUC value. Image-based machine learning models would be helpful for doctors to decide whether or not to perform a prostate biopsy. Future development of this technology will be further beneficial in making a diagnosis and treatment evaluation and patient prognosis.

## Data Availability

All relevant data are within the manuscript and its Supporting Information files.

## Acknowledgement

Technical assistance and critical advice are provided by the staff of the Department of Urology, Ciptomangunkusumo National Hospital.

